# Evaluation of serological tests for detecting SARS-CoV-2 antibodies: implementation in assessing post vaccination status

**DOI:** 10.1101/2021.04.27.21256205

**Authors:** Sally A. Mahmoud, Subhashini Ganesan, Shivaraj Naik, Safaa Bissar, Isra Zamil, Walid Abbas Zaher

## Abstract

**Background:** The anti-SARS-CoV-2 immunological assays have promising applications in the control and surveillance of the current COVID-19 pandemic. Therefore, large number of serological assays are developed in the commercial market to measure SARS-CoV-2 antibodies, which requires evaluation before their application in large scale.

**Objectives:** To evaluate the performances of commercially available serological assays for detecting SARS-CoV-2 antibodies.

**Methods:** The study compared the performances of six different methods for detection of antibodies against SARS-CoV-2 which includes (i) Genscript SARS-CoV-2 surrogate virus neutralization test kit [Test A] (ii) Diasorin - SARS-CoV-2 S1/S2 IgG detection [Test B] (iii) Alinity SARS-CoV-2 IgG II [Test C] (iv) Diasorin – SARS-CoV-2 TrimericS IgG [Test D] (v) Roche Elecsys Anti-SARS-CoV-2 – cobas [Test E] (vi) AESKULISA (AESKU Enzyme Linked Immunosorbent Assay) [Test F] against the gold standard Plaque Reduction Neutralization Test (PRNT).

**Results:** Test E had the highest sensitivity and Test A had the highest specificity The ROC for tests A, C, D and E showed optimum cut-offs that differed from the manufacturer’s recommendation. Test D had the best performance considering all the performance indicators with the highest agreement with the PRNT results. Parallel testing of test A with test D and test B had the optimum performance.

**Conclusion:** Serological assays that are commercially available are very promising and show good agreement with the standard PRNT results. Studies on large samples for optimization of the assay cut-off values and cost-effective evaluations on parallel testing methods are needed to make recommendations on these commercial assays.

**Importance:** Serological assays that are commercially available are very promising and this paper adds new knowledge about the optimization of these kits for evaluating post vaccination antibodies status. It highlights the positive and negative aspects of each of these assays in terms of sensitivity, specificity, positive and negative predictive values, and the agreement of results with the standard neutralization test. When serological assays are being used to assess post-vaccine immune status, a balance of all parameters needs to be considered rather than emphasizing only on high specificity. This is particularly relevant in the current situation where vaccination is happening around the globe, high sensitivity assays will result in reporting a lower percentage of false negative reports and avoids panic about lack of vaccine response. It is important that we understand the strengths and limitations of commercially available serological assays for better application of these tests to understand immune response and the duration of protection post vaccination.

## Background

The emergence of this global pandemic of COVID-19 has created an increased need for large scale PCR testing and serological assays. Serological testing has enormous applications in handling the current pandemic, it has both individual and population level practical applications that can facilitate pandemic response. At the individual level it can help differentiate recent and past infections of COVID-19, immune status post-vaccination to study the need for booster doses and identifying vaccine intervals. At the population level it helps study the epidemiology of SARS-CoV-2 pandemic, seroprevalence, and thereby help public health experts make recommendations on travel, social distancing, and the protective status of the population. [1] There are several serodiagnosis assay platforms that are being used for COVID-19 infections: the FDA has issued emergency authorization for ELISA, lateral flow immunoassay, and microsphere immunoassay. [2] These tests measure the antibody to nucleocapsid N protein antigens and antibodies binding to SARS-CoV-2 spike protein S, but not all spike-binding antibodies are functional or blocks viral infection, hence they do not indicate the functional measure of the antibody that inhibits SARS-CoV-2 infection. Ideally, tests should measure the neutralizing antibodies, which implicate protection from infection. The gold standard for measuring neutralizing antibodies is the plaque reduction neutralization test (PRNT). PRNT is however not practical for large scale as it requires skilled manpower, high-level biohazard security (BSL-3 level) and requires long turnaround time of five days. [3,4]

Therefore, to address this gap, a lot of commercial serological assays are developed and are now available in the market. A meta-analysis done on these serological assays for detecting antibodies against SARS-CoV-2 have shown that assays using the S antigen and testing IgG antibodies perform with better sensitivity than N antigen and Ig M based tests. [5]

It is important that we understand the strengths and limitations of commercially available serological assays for better application for these tests to understand immune response and the protection and duration of protection after vaccination. Hence this research tries to evaluate the various methods for detecting SARS-CoV-2 antibodies compared to the gold standard PRNT.

## Objective

To evaluate the performances of commercially available serological assays for detecting SARS-CoV-2 antibodies

## Methods

The study compared six different methods for detection of antibodies against SARS-CoV-2 detection post vaccination for COVID-19 against the gold standard Plaque Reduction Neutralization Test (PRNT). The six different methods are (i) Genscript SARS-CoV-2 surrogate virus neutralization test kit [Test A] (ii) Diasorin - SARS-CoV-2 S1/S2 IgG detection [Test B] (iii) Alinity SARS-CoV-2 IgG II [Test C] (iv) Diasorin – SARS-CoV-2 TrimericS IgG [Test D] (v) Roche Elecsys Anti-SARS-CoV-2 – cobas [Test E] (vi) AESKULISA (AESKU Enzyme Linked Immunosorbent Assay) [Test F]

The PRNT is a serological test which utilizes the ability of a specific antibody to neutralize a virus, in turn, preventing the virus from causing the formation of plaques in a cell monolayer. In this study, Vero E6 cells were grown to a confluent monolayer in a 6 well plate. The positive control was (Pooled Serum sample of Vaccinated and/or Covid-19 positive patient) and viral stock diluted in Dulbecco’s modification of Eagle medium (DMEM) was used as negative control.

Interpretation is based on 50% neutralization, which is the last dilution of serum capable of inhibiting 50% of the total plaques (virions). Negative control should have plaque count ≥ 50%, positive control should be plaque count ≤ 50% of negative control, and titer at which 50% reduction of plaques is there in comparison to the negative control is taken as the antibody titer. For positive PRNT results, the cut off for positive is 1:20 dilution.

The study was done on 125 samples, of which 69 stored de-identified excess serum samples collected from post-vaccination patients who requested post-vaccination antibody testing for antibody levels against SARS-CoV -2 Virus and 56 negative serum samples from non-vaccinated COVID-19 negative patients. Each individual sample was tested using the PRNT method and with all the six different serological assays, and the results of each method were compared to the gold standard reference method which is PRNT.

All serological assays were done based on manufacturers’ guidelines and recommendations. Table 1 shows the details of the various serological assays evaluated in this study. Test A and F were enzyme linked immunosorbent assay (ELISA) based tests, tests B,D and E were chemiluminescent immunoassay (CLIA) based tests and test D was based on chemiluminescent microparticle immunoassay (CMIA)

**Table 1:** Details of the six commercial serological assays.

| Test | Reagent/kit | Method | Manufacturer | Isotype | Target protein | Run time (minutes) |
| --- | --- | --- | --- | --- | --- | --- |
| A | cPass SARS-COV-2 surrogate virus neutralization test kit | ELISA | Genscript | IgG | RBD unit of S1 | 180 |
| B | Diasorin SARS-CoV-2 S1/S2 IgG detection | CLIA | Diasorin | IgG | S1/S2 | 35 |
| C | Alinity SARS-CoV-2 IgG II | CMIA | Abbott | IgG | RBD unit of S1 | 20 |
| D | SARS-CoV-2 TrimericS IgG | CLIA | Diasorin | IgG | RBD unit of S1 | 20 |
| E | Elecsys Anti-SARS-CoV-2 – cobas | CLIA | Roche | IgG & IgM | RBD unit of S1 | 20 |
| F | AESKULISA (AESKU Enzyme Linked Immunosorbent Assay) | ELISA | AESKU | IgG | S1 | 180 |

## Results

125 samples were used in this study and PRNT was done on all 125 samples. Due to a lack of sufficient sample volume, some of the serological assays were not performed for certain samples. The borderline/equivocal results that were above the cut-off values for positive reports were considered positive. Table 2 shows the number of samples tested using each assay and the results of each.

**Table 2:** Results of all six commercial serological assays.

| Assay | Positive | Negative | Total |
| --- | --- | --- | --- |
| Test A | 52 | 73 | 125 |
| Test B | 43 | 61 | 104 |
| Test C | 67 | 37 | 104 |
| Test D | 50 | 53 | 103 |
| Test E | 76 | 20 | 96 |
| Test F | 61 | 38 | 99 |

Test E had the highest sensitivity followed by test C and F. Test A had the highest specificity followed by test B and D. Test E though had the highest sensitivity had the lowest specificity compared to the other rapid tests as shown in Table 3.

**Table 3:** Performance indicators of the serological assays compared to PRNT.

| Assay | Sensitivity<br>(95% CI) | Specificity<br>(95% CI) | PPV<br>(95% CI) | NPV<br>(95% CI) | Overall<br>agreement<br>with PRNT<br>results (95% CI) |
| --- | --- | --- | --- | --- | --- |
| Test | 71.07 | 94.64 | 94.23 | 72.60 | 81.6 |
| A | (58.8–81.3) | (85.1-98.8) | (84.1-98.8) | (60.9-82.4) | (73.7-87.9) |
| Test | 75.47 | 94.12 | 93.02 | 78.69 | 84.61 |
| B | (61.7-86.2) | (83.8-98.8) | (80.9-98.5) | (66.3-88.1) | (76.2-90.9) |
| Test | 98.11 | 70.59 | 77.61 | 97.30 | 84.62 |
| C | (89.9-99.9) | (56.1-82.5) | (65.8-86.9) | (85.8-99.9) | (76.22-90.94) |
| Test | 84.91 | 90 | 90 | 84.91 | 87.37 |
| D | (72.4-93.2) | (78.1-96.6) | (78.1-96.6) | (72.4-93.2) | (79.4-93.1) |
| Test | 100 | 41.67 | 63.16 | 100 | 70.83 |
| E | (92.6-100) | (27.6-56.7) | (51.3-73.9) | (83.1-100) | (60.7-79.7) |
| Test | 92.31 | 72.34 | 78.69 | 89.47 | 82.82 |
| F | (81.5-97.9) | (57.4-84.4) | (66.3-88.1) | (75.2-97.1) | (73.9-89.7) |

When the high sensitivity tests E and C were followed by serial testing with tests A and B with high specificity, it improved specificity of the tests E and C with a lower sensitivity. [Figure 1] and [Figure 2]

**Figure 1:**
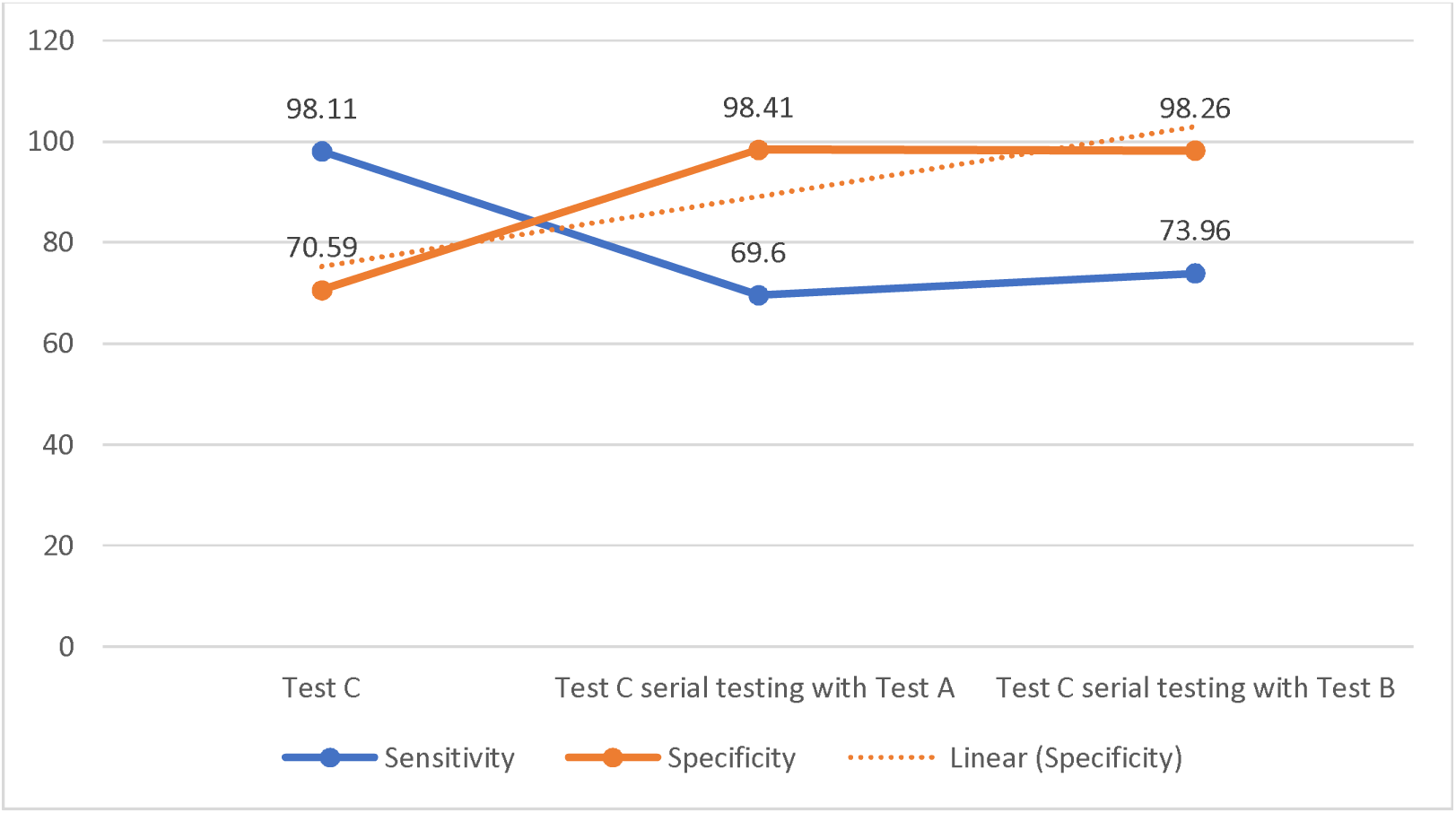
Serial testing of test C with test A and test B.

**Figure 2:**
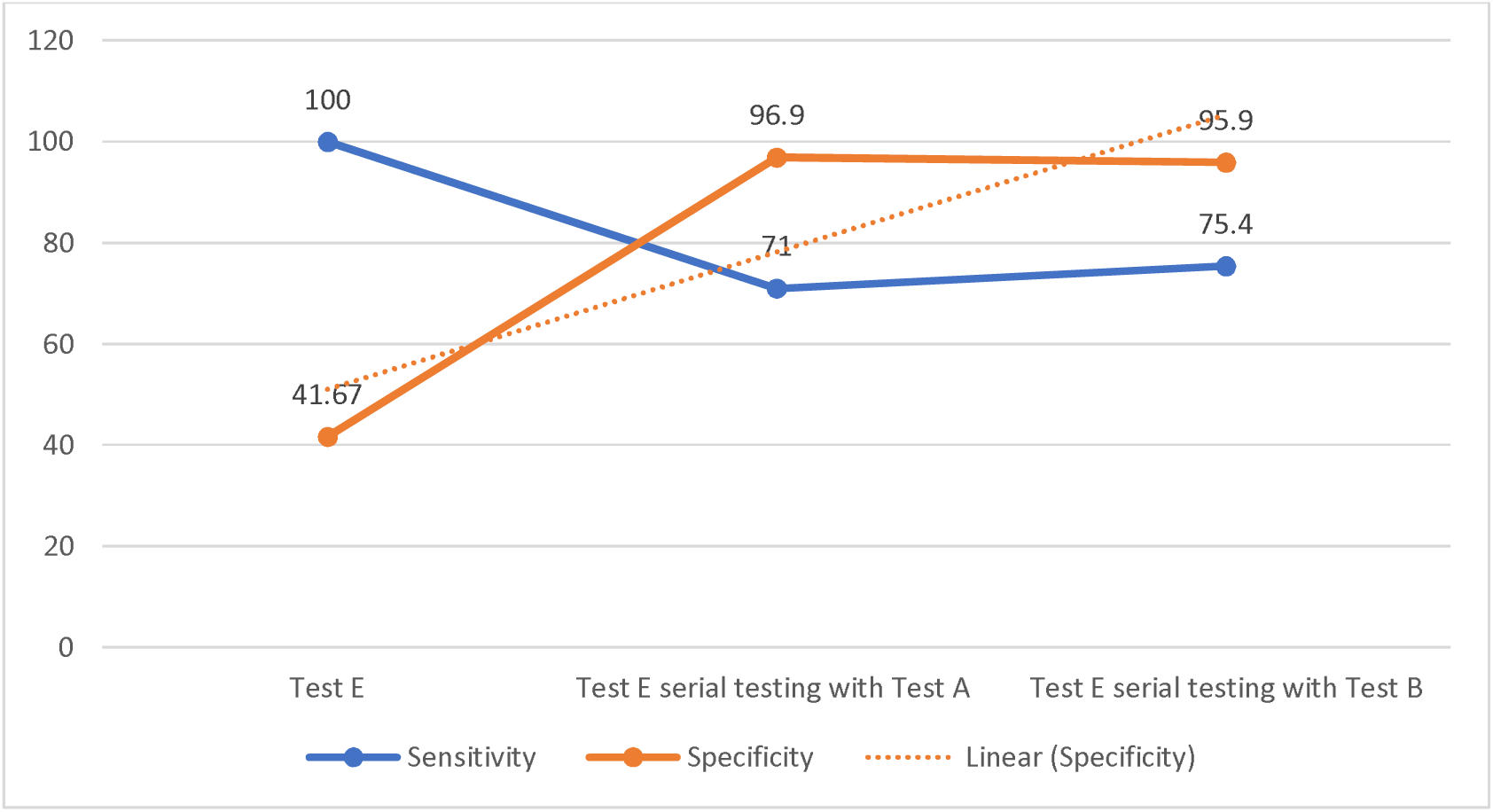
Serial testing of test E with test A and test B.

Also employing two tests with high specificity in parallel for example test A with test B or test D, improved the sensitivity to 92% and 95% respectively, with only a slight loss in specificity. [Table 4]

**Table 4:** Test A in parallel testing with other serological tests.

| Parallel testing |  | Sensitivity | Specificity |
| --- | --- | --- | --- |
| Test A | Test B | 92.86 | 89.07 |
|  | Test C | 99.46 | 66.80 |
|  | Test D | 95.63 | 85.17 |
|  | Test E | 100 | 39.72 |
|  | Test F | 97.77 | 68.46 |

When a functional test like test A is combined with a quantitative test in parallel and serial testing, it shows that test A in parallel testing with test B or test D has the optimum sensitivity and specificity. (Table 4 and Table 5) Test A in serial testing with test B shows the maximum specificity. (Table 5)

**Table 5:** Test A in serial testing with other serological tests.

| Serial testing |  | Sensitivity | Specificity |
| --- | --- | --- | --- |
| Test A | Test B | 53.53 | 99.69 |
|  | Test C | 69.65 | 98.41 |
|  | Test D | 60.27 | 99.46 |
|  | Test E | 71.0 | 96.9 |
|  | Test F | 65.54 | 98.52 |

ROC curve was plotted for all the serological assays and the area under the curve was largest for test D followed by test A and test B, showing the best performance to differentiate positive and negative results compared to the PRNT results. (Figure 3 and Table 6) Based on the ROC optimum cutoffs for the tests were estimated and these were similar to the manufacturer’s values for tests B and F. However, tests A, C, D and E showed different cut offs based on our ROC curves than the one recommended by the manufacturers. From the adapted ROC cut offs sensitivity, specificity, and overall agreement with the PRNT results were calculated. [Table 7]

**Table 6:** Area under the curve (AUC) for the serological assays.

| Antibody tests | AUC | 95% Confidence Interval |
| --- | --- | --- |
| Test A | 0.939 | (0.888 - 0.990) |
| Test B | 0.935 | (0.885 – 0.986) |
| Test C | 0.860 | (0.784 -0.936) |
| Test D | 0.953 | (0.912 - 0.994) |
| Test E | 0.839 | (0.751 - 0.927) |
| Test F | 0.928 | (0.871 - 0.984) |

**Table 7:** Optimum cut-offs based on the ROC curves and their performances.

|  | Cut offs | Sensitivity | Specificity | Overall agreement<br>with PRNT results |
| --- | --- | --- | --- | --- |
| Test A | Manufacturer's cut off – 30% | 71.0 | 94.23 | 81.6 |
|  | Based on ROC cut off – 20% | 92.31 | 87.5 | 90.4 |
| Test C | Manufacturer's cut off - 50 | 98.11 | 70.59 | 84.62 |
|  | Based on ROC cut off - 150 | 83.01 | 88.23 | 85.57 |
| Test D | Manufacturer's cut off - 33.8 | 84.91 | 90 | 87.37 |
|  | Based on ROC cut off - 40 | 84.90 | 96.0 | 90.29 |
| Test E | Manufacturer's cut off - 0.8 | 100 | 41.67 | 70.83 |
|  | Based on ROC cut off - 5 | 93.75 | 70.83 | 82.29 |

\* Tests B and F showed ROC based cut offs similar to the manufacturer's values.

**Figure 3:**
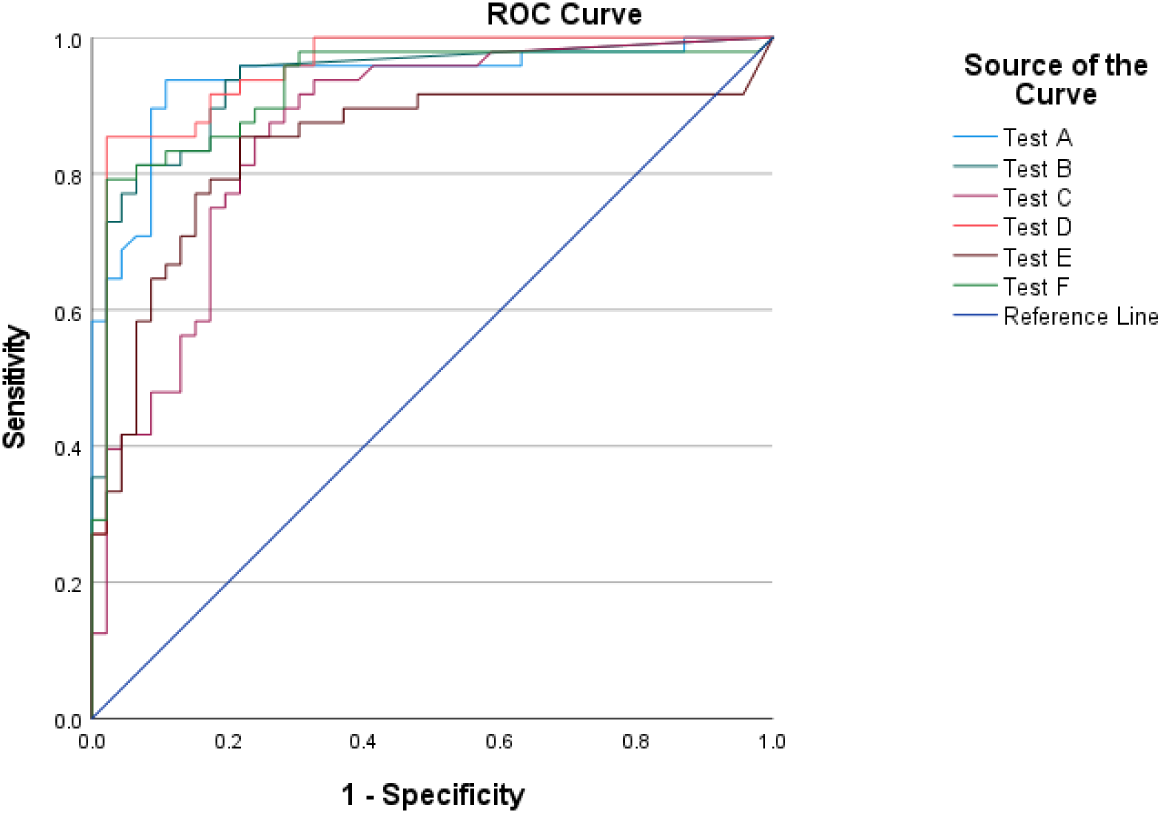
ROC curves for the serological assays.

For test A, the manufacturers had an initial cut-off of 20% inhibition, which was changed due to the FDA recommendations to 30% to increase specificity. However, comparing test A results with the PRNT reports, showed 20% as optimum cut-off. The scatter plot of the distribution of % inhibition of test A based on the PRNT reports show the number of positive cases missed, with the increase in cut-off values by 10%. [Figure 4] Test A showed higher sensitivity and overall agreement with PRNT when 20% inhibition was used as a cut-off then the recommended 30%. [Figure 5]

**Figure 4:**
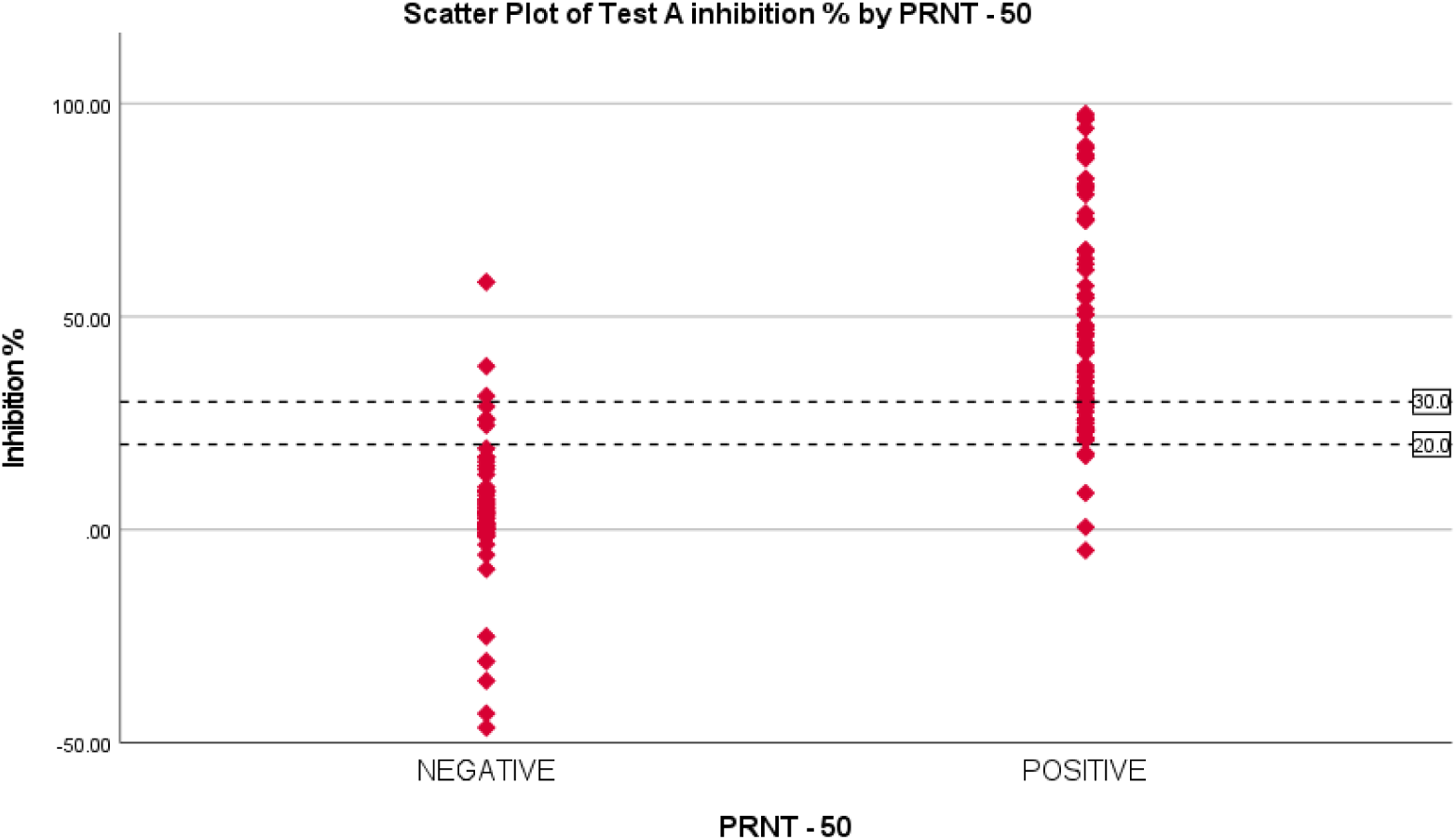
Scatter plot of test A % inhibition based on results of the PRNT.

**Figure 5:**
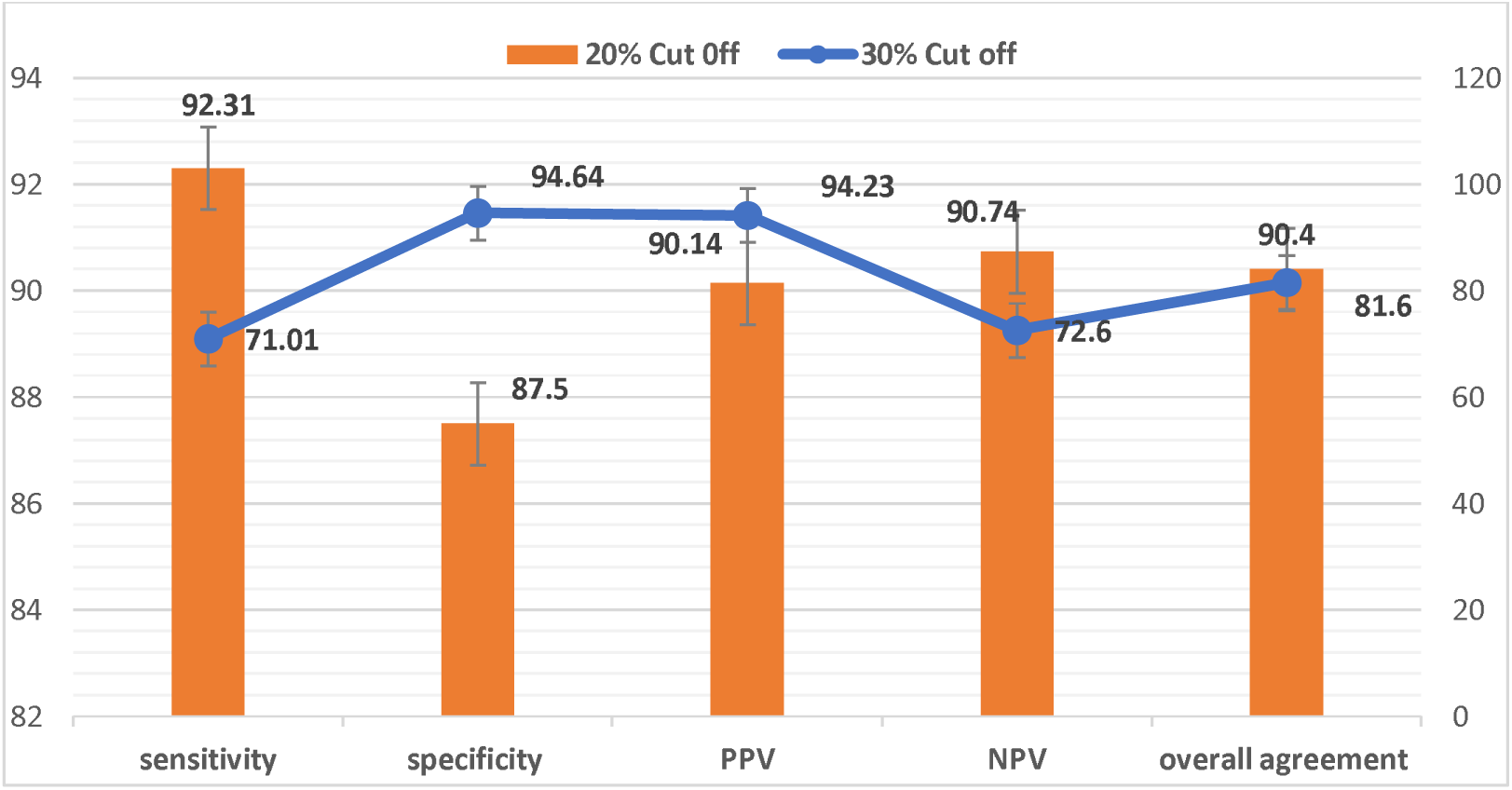
Comparison of the test A performance based on two different cut-off values.

## Discussion

This study compared six immunoassays for detection of IgG antibodies against SARS-CoV-2 against the standard viral neutralization test PRNT.

In this study, tests A and B had the lowest sensitivity but had a high specificity and PPV. But studies that evaluated test A had demonstrated test A to have high sensitivity [6,7]. However, this might be because all these studies used cutoff as 20% inhibition, which was later changed according to FDA recommendation to 30%. [8] A study that compared test A to PRNT similar to our study used 30% as cutoff and showed that sensitivity ranged from 77% - 100% and specificity was 95% - 100% [9], which was similar to our study results when 30% cutoff was used.

The ROC curve adapted cutoff for test A was around 20% inhibition and when 20% was used as a cutoff for test A the sensitivity increased to 92.31% and the overall agreement with the PRNT results was better (90.4%) suggesting 20% inhibition as a more optimum cut off for the test.

Another study that evaluated test A suggested that the test might require specific cutoffs with respect to ethnic, geographical background and the prevalence of COVID-19 infections. The study also showed that introduction of an equivocal range with repeat testing within the range of 18 – 22% can reduce the false positive results.[10]

Based on the ROC threshold values, tests D and E did not correlate with the manufacturers cut-off. Raising the cut-offs for tests D and E as per the ROC values showed increased specificity and agreement with the PRNT results without much change in sensitivity. However, for test C raising the cut-off increased specificity with a huge drop in sensitivity, but the agreement with the PRNT results were better.

Studies suggest revision of cut-off values provided by manufacturers, as most of the assay validation are done on a small sample size and among specific ethnic or regional group.[11] Therefore more evaluation studies and optimum cut-offs need to be defined before these serological assays are used in large scale to evaluate vaccination status of the population.

Test E had the highest sensitivity in this study as reported in similar studies, but these studies have also reported high specificity, which our study did not find. [12,13]

The sensitivity of test B was similar to that reported in other studies [12]. Studies evaluating C and E demonstrated higher specificity of test C and test E compared to our study. [14] A meta-analysis on antibody tests for SARS-CoV-2 showed that tests using ELISA and CLIA-based methods performed better [5]. In our study test A and F are based on ELISA and tests B, D and E were CLIA-based tests. While tests D, E and F showed higher sensitivities, the sensitivities of test A and B were low.

Test C was evaluated by a study which compared antibodies in post-vaccination patients compared with pre-pandemic serum samples and claims high sensitivity and a specificity of 100%. While our study showed that the sensitivity of test C was high, the specificity was low compared to this study. [15] This might be because these studies have not compared the assays with the PRNT method, but with the RT-PCR assay validation.

Test D had good sensitivity and specificity with the highest agreement with the PRNT results, as also supported by another that evaluated test D. [16]

In addition, when a functional test like test A was tested with quantitative tests in parallel, it is observed that the overall sensitivity increases. An optimum sensitivity and specificity are achieved when test A is done in parallel with test B. This kind of combination of two antibody tests are being studied and it shows that it increases the ability to capture the positive results. [17]

The CDC recommends serological tests with high sensitivity and specificity and tests detecting IgG or both IgG and IgM. This is because currently serological tests are recommended by CDC only for identifying persons with previous infections or to identify resolving infections and for better understanding the epidemiology of SARS-CoV-2. [18] However, these antibody tests help us understand the development of immune response and the longevity of the antibodies developed post-vaccination for COVID-19. This surveillance becomes essential to identify vaccine efficiency and make recommendation on booster doses and the intervals for vaccination.

The WHO, in collaboration with Coalition for Epidemic Preparedness Innovations (CEPI), and the National Institute for Biological Standards and Control (NIBSC), has come up with the International standards for anti-SARS-CoV-2 immunoglobulins. This is very crucial for a standard comparison as vaccine developers have been using various immunoassays with different measuring units making the comparison of immunogenicity difficult. Hence with these recommendations future studies can make the comparison of immunogenicity more standardized and will be less challenging. [19]

Most COVID-19 vaccinations use the S protein or S-domains as immunogen target. [20] Therefore these serological assays that target antibodies against S protein and the RBD directed IgG detection serve as a good candidate for evaluating vaccine response. Therefore, keeping this in mind, when serological assays are being used to assess post-vaccine immune status, a balance of all parameters needs to be considered rather than emphasizing more on only high specificity. This is particularly relevant in the current situation where vaccination is happening around the globe and the percentage of vaccination is increasing, high sensitivity assays will result in reporting a lower percentage of false negative reports. This will help assess the immune status and avoids panic about lack of vaccine response. Thus, recommendations based on a balance of all these parameters are needed.

## Strengths

Most evaluation studies compare the serological assays with RT-PCR reports. This study is one of its kind that compares commercial serological assays using the same serum sample and by evaluating the results with the gold standard PRNT reports. Thus, this study reduces the biases and provides a standard comparison.

## Limitations

The number of samples were limited and additional parameters like days post vaccination and the type of vaccine were not taken into consideration in this study, which could have provided further insights on the serological assay performances.

## Conclusion

Serological assays that are commercially available are very promising and show good agreement with the standard PRNT results. We recommend further studies of these serological assays with large number of samples, to understand more about the performance of these assays. Moreover, performing two tests in parallel testing improves the sensitivity and a better alternative to conventional PRNT, however cost-effective evaluations are needed to recommend these. We suggest optimization of the cut offs values for these serological assays considering the prevalence, ethnic and geographical variations. Recommendations based on the balance of all performance indicators rather than just specificity will help in the application of these serological assays in assessing post-vaccination status.

## Data Availability

The data is available with the corresponding author, Dr. Sally, Director of Biogenix G42 lab and will be produced on request

## Ethics approval and consent to participate

The Ethics approval was obtained from Department of Health (DOH) Institutional review board (IRB), Abu Dhabi. All methods were carried out in accordance with relevant guidelines and regulations.

## Informed consent statement

Informed consent was waived off by the Department of Health (DOH) Institutional review board (IRB), Abu Dhabi.

